# Predicting Conversion from SCD to MCI: A Machine Learning Study

**DOI:** 10.64898/2026.09.04.26362240

**Authors:** Farooq Kamal, Amelie Metz, Katherine Chadwick, Roqaie Moqadam, Cassandra Morrison, Mahsa Dadar, Alzheimer’s Disease Neuroimaging Initiative, Consortium for the Early Identification of Alzheimer’s Disease-Quebec (CIMA-Q), the PREVENT-AD Research Group

**Author notes:** Corresponding authors: Farooq Kamal, Mahsa Dadar Douglas Mental Health University Institute, Montreal, Quebec, Canada H4H 1R3. Data used in preparation of this article were obtained from the Alzheimer’s Disease Neuroimaging Initiative (ADNI) database (adni.loni.usc.edu). As such, the investigators within the ADNI contributed to the design and implementation of ADNI and/or provided data but did not participate in analysis or writing of this report. A complete listing of ADNI investigators can be found at: http://adni.loni.usc.edu/wp-content/uploads/how_to_apply/ADNI_Acknowledgement_List.pdf. Data used in preparation of this article were obtained from the PRe-symptomatic EValuation of Experimental or Novel Treatments for Alzheimer’s Disease (PREVENT-AD) program, data release 8.1 (https://www.centrestopad.com/). Data used in the preparation of this article were obtained from the Consortium for the early identification of Alzheimer’s disease – Quebec (CIMA-Q; cima-q.ca). A list of researchers involved in the design of CIMA-Q can be found on the cima-q.ca website. These researchers contributed to the establishment of protocols, the implementation of the research infrastructure, the recruitment and follow-up of participants, the obtaining of data, the maintenance of biological and ex-vivo samples, and certain derived data.

## Abstract

**BACKGROUND:** Subjective cognitive decline (SCD) may precede mild cognitive impairment (MCI), but not all individuals with SCD progress to MCI. Identifying which individuals are most likely to convert and over what time frame remains an important goal in Alzheimer’s disease research. MRI measures of white matter hyperintensity (WMH) burden and gray matter (GM) atrophy may improve prediction beyond demographic and cognitive predictors, but their incremental value across different time intervals has not been established.

**METHODS:** Data were obtained from four longitudinal cohorts (ADNI, NACC, CIMA-Q, and PREVENT-AD). A total of 1,352 participants with SCD at baseline were included. Machine learning models (logistic regression, random forest, XGBoost) were used to predict conversion from SCD to MCI at 2 years, 3 years, 4 years, and 5-year horizons. Four feature sets were compared: base (age, sex, education, APOE4, hypertension), base and cognition (adding MoCA and Trail Making Test Part B), base and MRI (adding regional WMH and GM volumes), and combined (all features).

**RESULTS:** The base and MRI set achieved the highest Area Under the Curve (AUC) at the 3-year (0.900), 4-year (0.879), and 5-year (0.923) horizons. The combined set achieved the highest AUC at the 2-year horizon only (0.884). MRI features produced larger AUC gains over the base model than cognitive features at the 3, 4, 5-year horizons. Parietal WMH was the most frequently selected MRI predictor.

**CONCLUSIONS:** MRI features, particularly regional WMH and GM volumes, provided greater predictive value than cognitive features at longer prediction horizons. A select number of regional MRI features predicted SCD to MCI conversion with high accuracy up to 5 years in advance.

## Introduction

Alzheimer’s disease (AD) is a progressive neurodegenerative disorder that is characterized by the accumulation of amyloid-beta plaques and neurofibrillary tau tangles [1]. These pathological changes begin years before the onset of clinical symptoms and cognitive decline [2]. Additional brain changes, including gray matter (GM) atrophy and white matter hyperintensity (WMH) burden, also contribute to cognitive decline [3, 4]. WMHs are markers of cerebrovascular injury and may exhibit region-specific patterns of accumulation across the brain as the disease progresses [5, 6]. Because there are currently limited treatments that can reliably halt or reverse disease progression, and available treatments (e.g., lecanemab) are more effective when initiated at earlier stages, identifying individuals at increased risk before the onset of cognitive decline remains an important goal in AD research.

Subjective cognitive decline (SCD) is considered the earliest symptomatic stage along the AD continuum and may precede mild cognitive impairment (MCI) [7, 9]. SCD refers to self-reported decline in cognition despite performance that remains within normal limits on standardized cognitive tests [7, 8]. SCD is common in older adults, affecting approximately one quarter of adults aged 60 years and older [10]. Individuals with SCD have been shown to have lower baseline cognition and faster cognitive decline across several domains compared to individuals without SCD [11]. Research has also shown greater WMH burden in SCD, particularly in temporal and parietal regions [12]. Post-mortem studies suggest that individuals with high AD pathology load reported SCD several years prior to death [13]. Taken together, these findings suggest that, even while cognitive tests remain normal, SCD may reflect early brain and cognitive changes that are relevant to later clinical decline.

Not all individuals with SCD progress to MCI or dementia. Annual conversion rates from SCD to MCI have been estimated at 6.7%, while annual conversion to dementia has been estimated at 2.3% [14]. Over a four-year period, approximately 25% of individuals with SCD progress to MCI [14]. This risk is twice as high compared to older adults without SCD. Individuals who are at a higher risk of preclinical AD compared to more general SCD, meet the SCD plus criteria which includes factors such as memory complaints, onset of decline within the past five years, age of at least 60 years, and associated concern or worry [7, 15]. Therefore, an important unresolved question is not simply whether SCD is associated with later decline, but which individuals with SCD are most likely to convert and over what timeframe [16].

Several demographic, genetic, cognitive, and vascular factors have been associated with conversion from SCD to MCI, including age, APOE4, hypertension, and education [17, 28]. In older women, the combination of SCD and an APOE4 allele shortens the time to MCI or dementia conversion, and women with SCD decline more rapidly than men across multiple cognitive domains [18, 19]. Lower global cognition, poorer verbal learning, and slower executive processing also predict later SCD conversion even in individuals who appear cognitively normal at baseline [17, 20]. Thus, subtle cognitive changes appear to add predictive value beyond demographic factors alone [17, 21].

Neuroimaging markers can refine risk estimates for conversion from SCD to MCI. Higher WMH burden has been reported to predict cognitive decline and MCI [12, 22], and may partly account for the association between SCD and later cognitive impairment [12, 22]. Research has shown that vascular injury accumulates unevenly across the brain, thus regional WMH burden compared to only total WMH are likely to be informative for prediction [4, 23]. In addition to WMH burden, GM atrophy in temporal, parietal, and hippocampal regions has been associated with progression to MCI [24, 25]. Studies that combine MRI with clinical measures generally outperform those based solely on clinical measures [13]. However, the additional value of MRI after demographic, genetic, vascular, and cognitive factors are considered remains uncertain. The present study addresses this question directly by comparing different combinations of predictors.

Machine learning (ML) approaches may be useful to address this gap in the literature because they can model complex relationships among predictors that may not be captured by traditional linear approaches [13, 26]. One study using a nonlinear XGBoost classifier found that including age, depression scores, and GM predicted conversion from SCD to MCI or AD with high predictive performance in a selected train/test split (AUC=0.96) [27]. Not only is such research limited but they also rely on very small sample sizes, smaller number of predictors, single follow-up windows or have not directly compared the contribution of cognitive and neuroimaging variables within the same sample. It remains unclear whether different types of predictors are more useful at shorter versus longer prediction horizons that is, the number of years between baseline assessment and the point at which conversion occurs.

The present study developed and evaluated ML models to predict conversion from SCD to MCI across 2, 3, 4, and 5-year horizons. To overcome the limited sample sizes of previous studies, data from four longitudinal cohorts were harmonized and merged into a larger sample. Four sets of predictors were compared: (1) a base model including demographic, genetics, and vascular information; (2) a base plus cognition model that also included cognitive tests; (3) a base plus MRI model that also included regional WMH and GM volumes; and (4) a combined model including all predictors. This approach examines whether cognitive and MRI features improve prediction beyond standard demographic, genetic, and vascular risk factors. We hypothesized that the combined model would show the strongest performance, and MRI features would provide greater incremental benefit at longer prediction horizons.

## Methods

### Datasets and Participants

Data were obtained from four independent longitudinal cohorts: the Alzheimer’s Disease Neuroimaging Initiative (ADNI), the National Alzheimer’s Coordinating Center (NACC), the Consortium for the Early Identification of Alzheimer’s Disease-Quebec (CIMA-Q), and the Pre-symptomatic Evaluation of Experimental or Novel Treatments for Alzheimer’s Disease (PREVENT-AD). Participants with unstable diagnostic trajectories such as those who converted to MCI and later reverted to SCD were excluded. The combined baseline sample included 1,352 participants with SCD. Of these, 195 converted to MCI during the follow-up period and 1,157 remained stable with SCD and no cognitive decline.

### Participants in ADNI

Data were obtained from the ADNI database (adni.loni.usc.edu). ADNI was launched in 2003 as a public-private partnership led by Principal Investigator Michael W. Weiner, MD. The initiative’s primary objective is to determine whether serial MRI, positron emission tomography, other biological markers, and clinical and neuropsychological assessments can be combined to measure the progression of MCI and early AD. Participants were selected based on availability of neuroimaging data and clinical assessments [29, 30]. Detailed inclusion and exclusion criteria are available at www.adni-info.org. Participants were classified as having SCD if they met ADNI criteria for subjective memory concern and had available MRI data from which regional WMH and GM volumes could be extracted. SMC is defined by a self-reported memory concern quantified on the Cognitive Change Index (total score above 16 on the first 12 items), no informant-corroborated concern, a Clinical Dementia Rating of 0, and normal performance on the Mini-Mental State Examination and Wechsler Logical Memory II delayed recall. A total of 270 participants met these criteria. Conversion was defined as the first subsequent visit at which the participant received a diagnosis of MCI. ADNI classifies MCI by a subjective memory concern, an MMSE of 24 to 30, a CDR of 0.5 with a memory box score of at least 0.5, objective memory impairment on Wechsler Logical Memory II delayed recall scored below cutoffs adjusted for education (≤8 for 16 or more years, ≤4 for 8 to 15 years, ≤2 for 0 to 7 years), and preserved functional independence [30].

### Participants in NACC

Data were obtained from the NACC database (https://naccdata.org/), including the Uniform Data Set (UDS) and MRI Data Set [31–33]. Participants were included if they had available MRI data from which regional WMH and GM volumes could be extracted. SCD was defined as the first visit where cognitive status (NACCUDSD) was classified as normal cognition and the participant reported subjective cognitive decline (DECSUB = 1). A total of 749 participants met these criteria. Conversion was defined as the first subsequent visit at which NACCUDSD reached 3 (MCI). NACC assigns MCI using the NIA-AA core clinical criteria: concern about a change in cognition reported by the subject, co-participant, or clinician; impairment in one or more cognitive domains; and largely preserved functional independence [34].

### Participants in CIMA-Q

Data were obtained from the CIMA-Q database [35]. CIMA-Q is a multicenter longitudinal cohort study that recruits and tracks disease progression in individuals at risk of dementia across Quebec. SCD was defined as the first visit at which the participant answered “Yes, this worries me” to the question “Do you feel like your memory is becoming worse?” Participants classified with SCD also had a MoCA score of at least 26, a CDR global score of 0, and normal education-adjusted Logical Memory performance. A total of 130 participants met these criteria. Conversion was defined as the first subsequent visit with a diagnosis of MCI. MCI classifications required a reported worsening of memory, regardless of whether it caused worry, a CDR global score of 0.5, and fulfilment of the National Institute on Aging–Alzheimer’s Association clinical core criteria for MCI [34].

### Participants in PREVENT-AD

Data were obtained from the PREVENT-AD database [36]. PREVENT-AD is a longitudinal study of cognitively unimpaired older adults with a parental or multiple-sibling history of AD, based at the Douglas Mental Health University Institute in Montreal. SCD was defined as the first questionnaire visit at which the participant endorsed worsening memory (scd_memory_becoming_worse). A total of 203 participants met these criteria. Conversion was defined as the first recorded MCI visit date after baseline. Conversion was defined as the first recorded initial_MCI_visit_date occurring after SCD baseline. PREVENT-AD initially screened cognition using the MoCA and CDR. When cognitive status was uncertain, typically because of a MoCA score of approximately 26 or lower or a CDR score above 0, participants received a comprehensive 2.5-hour assessment by a certified neuropsychologist. During follow-up, performance more than one standard deviation below the RBANS mean in two cognitive domains also triggered a comprehensive neuropsychological evaluation. Participants judged to have probable MCI following this evaluation had their conversion date documented in the repository.

Across all four datasets, SCD baseline was defined as the first visit meeting each dataset’s SCD criteria. Conversion was defined as the first subsequent visit meeting each dataset’s MCI criteria. Because the cohorts operationalize these constructs differently, ranging from questionnaire-based classification in ADNI and PREVENT-AD to clinician diagnosis in NACC and CIMA-Q, definitions were harmonized to a common SCD and MCI labeling scheme across cohorts. The primary outcome was conversion to MCI at any point after baseline. Participants who did not convert during follow-up were classified as stable. Participants who reverted to cognitively healthy status after their initial SCD diagnosis were excluded. Clinical variables including age, sex, education, APOE4 genotype, hypertension, Montreal Cognitive Assessment (MoCA), and Trail Making Test Part B (TMT-B) were merged to each participant’s baseline visit using the closest available assessment.

### MRI Processing

All T1-weighted (T1w) scans were preprocessed using a standard pipeline [37] that includes noise reduction [38], intensity inhomogeneity correction [39], and intensity normalization into range [0–100]. The preprocessed images were linearly registered (9 parameters: 3 translation, 3 rotation, and 3 scaling) [40] to the MNI-ICBM152-2009c average template [41], and all registrations underwent visual quality control.

### WMH measurements

WMH segmentation was performed on T1w images using a validated automated technique [42]. The technique extracts a set of location (spatial priors) and intensity (distribution histograms) features and uses them in combination with a random forest classifier to detect WMHs. This approach has been validated in multi-center studies [37,43] and across the ADNI and NACC cohorts [6]. Automatic segmentation was completed using only T1w contrasts, because FLAIR availability and resolution varied across datasets and sub-studies. We have previously shown that our T1w-based WMH volumes correlate strongly with FLAIR-based measurements [44]. The quality of all WMH segmentations was visually assessed (MD, KC, and AM), and cases that failed quality control were excluded. WMH load was defined as the volume of all voxels identified as WMH in standard space (in mm^3^) and were thus normalized for head size. Regional (frontal, temporal, parietal, and occipital) and total WMH were calculated using the Hammers Atlas [42,44]. Regional WMH values were averaged across hemispheres to obtain one measure per region.

### Gray matter measurements

Regional GM measures were quantified using deformation-based morphometry (DBM), which uses deformations estimated based on nonlinear registrations to detect local morphological differences [45,46]. For each lobe, left and right GM values were averaged within each subregion, followed by an equal-weighted average across subregions. Frontal lobe regions included rostral middle frontal, caudal middle frontal, superior frontal, lateral and medial orbitofrontal, pars opercularis, pars triangularis, pars orbitalis, rostral and caudal anterior cingulate, precentral, and insula. Parietal lobe regions included superior parietal, inferior parietal, supramarginal, postcentral, precuneus, paracentral, posterior cingulate, and isthmus cingulate. Temporal lobe regions included superior temporal, middle temporal, inferior temporal, transverse temporal, fusiform, entorhinal, parahippocampal, hippocampus, and amygdala. Occipital lobe regions included lateral occipital, pericalcarine, cuneus, and lingual [47]. Total GM was computed as the mean of the four lobar values.

### Clinical and Demographic Variables

Age at baseline, sex, education, APOE4 carrier status, and hypertension were recorded for all participants. These factors were consistently present across datasets. APOE4 carrier status was coded as binary (1 = one or more e4 alleles, 0 = no e4 alleles). APOE coding differed across datasets and all codings were harmonized to the binary carrier variable. Hypertension was also harmonized to binary (1 = hypertension, 0 = normal). Cognitive measures included the MoCA total score and TMT-B completion time (seconds) as they were consistently available across all datasets.

Demographic differences between groups were assessed using independent-samples *t*-tests for continuous variables including age and education. Chi-square (χ²) tests were used to assess categorical variables such as sex and APOE carrier status. APOE4 status was coded by comparing participants with one or two APOE ε4 alleles with those who had none. Continuous variables were standardized across datasets.

### Prediction Outcome and Horizon Definition

The prediction outcome was binary (1 = converted from SCD to MCI within the specified horizon, 0 = remained stable). Four prediction horizons were evaluated: 2, 3, 4, and 5 years. For a given horizon α, participants were retained if they either (a) converted to MCI within horizon/years or (b) had at least α horizon/years of follow-up without conversion. Participants who converted after the horizon window were excluded from that horizon’s analysis to avoid misclassifying late converters as stable. Participants with no follow-up to determine their status at a given horizon were also excluded. At longer horizons, more participants met the conversion criterion, and the number of converters increased from 43 at α = 2 years to 106 at α = 5 years. The same participant could contribute to more than one horizon. The samples across horizons were therefore overlapping.

### Feature Sets

Four predefined feature sets were compared at each horizon. The base set included baseline age, sex, education, APOE4 carrier status, and hypertension (5 features). This base set served as the reference model. The base and cognition set added MoCA and TMT-B time to the base set (for a total of 7 features). The base and MRI set added five regional and total WMH (frontal, temporal, parietal, occipital, total) and five regional and total GM (frontal, temporal, parietal, occipital, total) to the base set, for a total of 15 features. The combined set included all base, cognition, and MRI features (17 features).

### Machine Learning Pipeline

Classification was performed using a nested repeated cross-validation framework. Three classifiers were evaluated: logistic regression, random forest, and XGBoost. Figure 1 summarizes the machine learning pipeline. Analyses were performed in Python 3.11.

**Figure 1.**
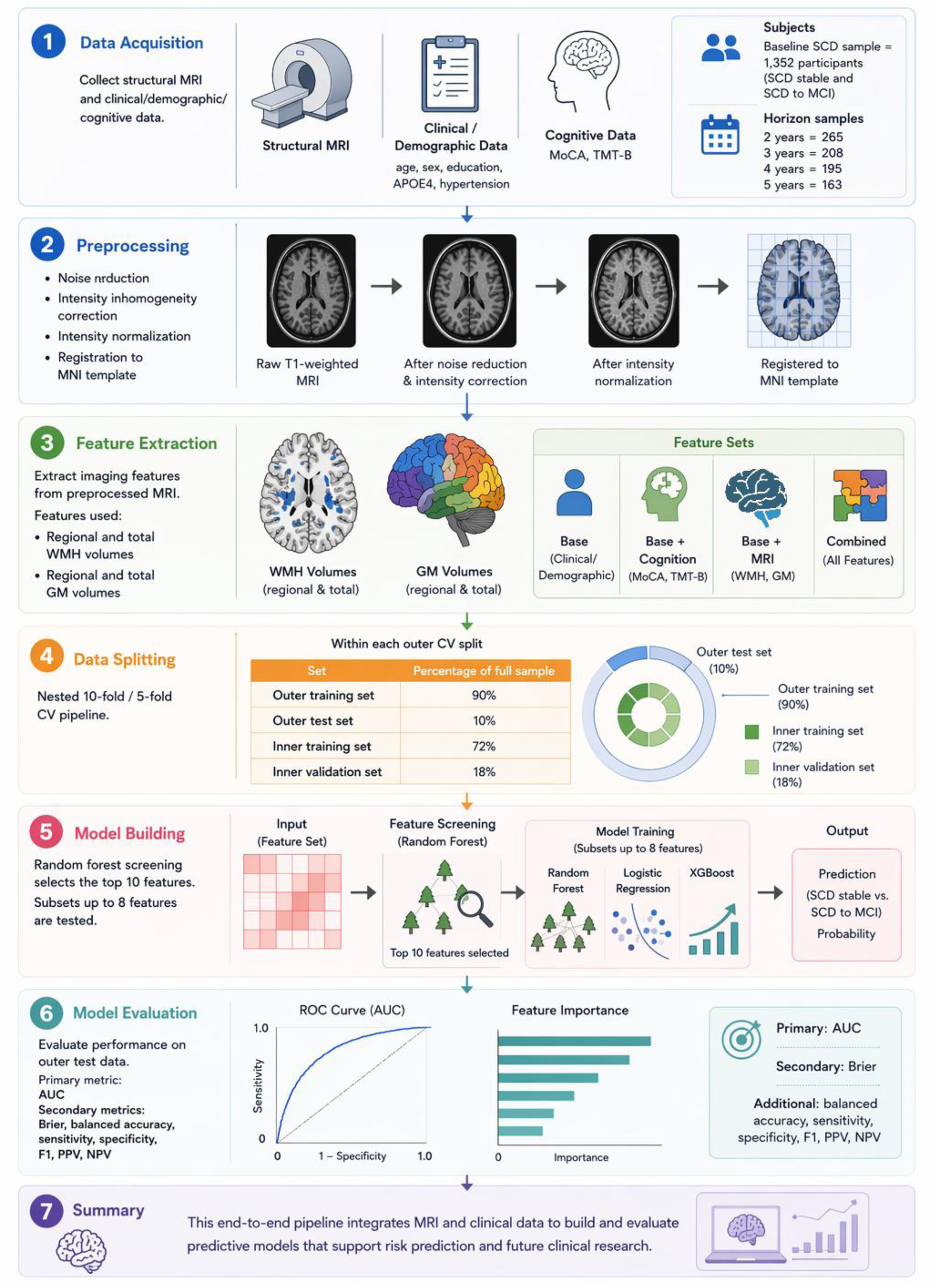
Machine learning methodology for predicting SCD to MCI conversion. (1) MRI, clinical/demographic, and cognitive data were obtained from four cohorts; (2) T1-weighted MRI scans were preprocessed and registered to the MNI template; (3) Regional and total WMH and GM were extracted and combined with clinical and cognitive predictors; (4) Four feature sets were evaluated across 2, 3, 4, and 5 year horizons; (5) Nested cross-validation was used for feature screening, subset selection, and model training; (6) Models were evaluated using AUC as the primary metric and Brier score as the secondary metric; (7) Final outputs included model performance, feature selection, and predicted conversion probabilities.

**Figure 2.**
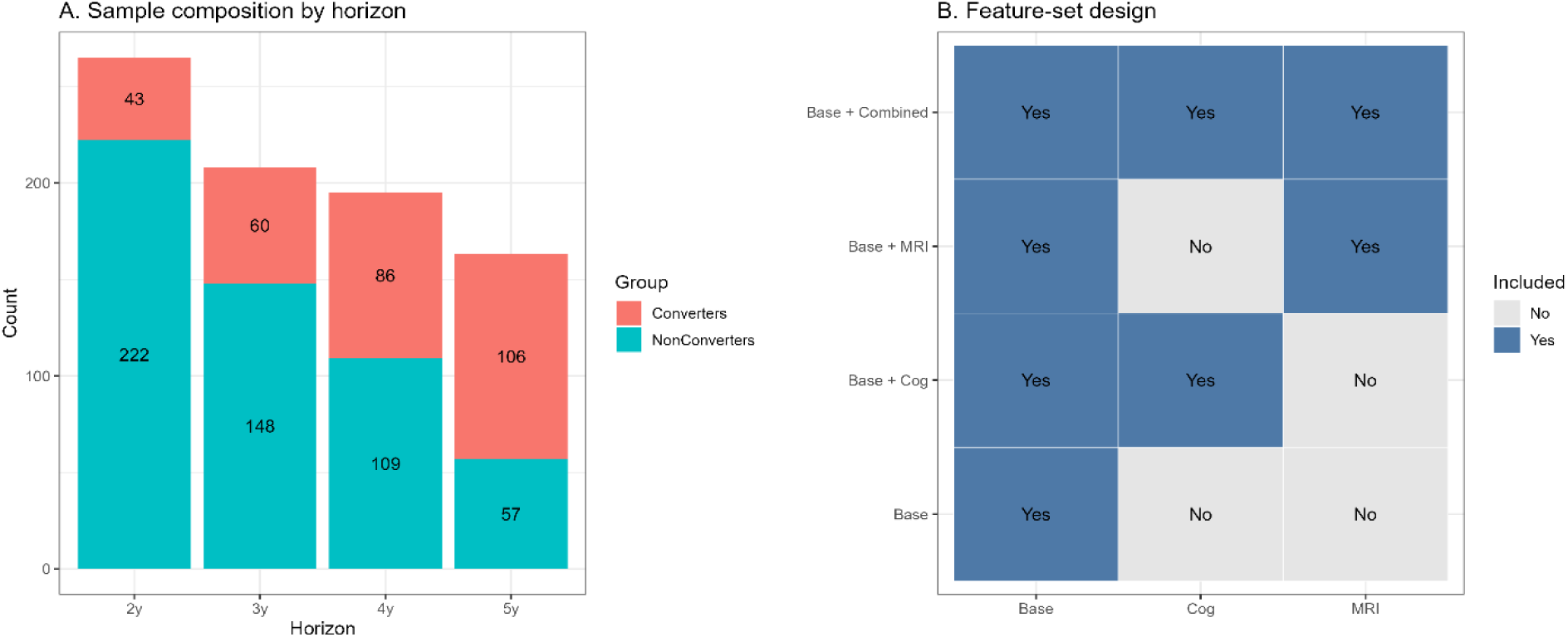
Study design and sample composition. Panel A shows the number of converters and non-converters included at each prediction horizon. Panel B shows which feature groups were included in each feature set.

### Nested cross-validation

The evaluation framework used an outer loop of 20 repeats of 10-fold stratified cross-validation. Each repeat used a different random partition to assess variability across splits. Within each outer training fold, an inner loop of 5-fold stratified cross-validation was used for model and feature subset selection. Inside each inner fold, a random forest-based screening step selected the top 10 most important features from the available pool. All combinatorial subsets of these screened features (up to size 8) were then evaluated. The best model and feature subset combination was selected based on the inner-fold area under the receiver operating characteristic curve (AUC, primary) and Brier score (secondary), measures of predictive performance. The selected model and subset were retrained on the full outer training fold and evaluated on the held-out outer test fold [48]. Larger subsets were retained only when they improved validation performance. Smaller subsets were preferred when performance was comparable, as complex models tend to generalize less well when sample sizes are limited [49–51].

### Final model selection

Across all 20 outer repeats, the model and feature subset combination most frequently selected was identified. This final model was re-evaluated in a single 10-fold stratified cross-validation to produce out of fold (OOF) predictions for the full sample. OOF predictions refer to predicted probabilities generated for each person when that person was held out from model training, providing an estimate of performance on unseen data.

### Evaluation metrics

The primary performance metric was the AUC. Secondary metrics included Brier score, balanced accuracy, sensitivity, specificity, and F1 score. Performance was reported as mean and 95% confidence intervals (2.5^th^ and 97.5^th^ percentiles) across the 20 outer repeats (OOF).

## Results

### Demographic differences across datasets

The full sample included 1,352 participants from ADNI (*N*=270), NACC (*N*=749), CIMA-Q (*N*=130), and PREVENT-AD (*N*=203). Age differed across datasets (*F*=27.34, *p*<.001). CIMA-Q participants were older than those from ADNI and NACC (*t*>2.29, *p*<.048), whereas PREVENT-AD participants were younger than participants from all three other datasets (*t*>9.70, all *p*<.001). Age did not differ between ADNI and NACC (*p*>.05).

Education also differed across datasets (*F*=11.18, *p*<.001). ADNI participants had more years of education than participants from NACC, CIMA-Q, and PREVENT-AD (*t*=3.83–6.45, all *p*<.001). Education did not differ among NACC, CIMA-Q, and PREVENT-AD (all *p*>.05). Sex differed across datasets (χ*²*=10.93, p=.012). PREVENT-AD had a lower proportion of male participants than ADNI and NACC (χ²=7.01 and 7.06, respectively; both *p*=.047). The remaining pairwise differences in sex were not significant. APOE4 carrier status also differed across datasets (χ²=9.78, *p*=.020), with a lower proportion of carriers in CIMA-Q than in PREVENT-AD (χ²=9.41, *p*=.013). No other pairwise differences in APOE4 status were significant. The prevalence of hypertension differed across datasets (χ²=39.58, *p*<.001). PREVENT-AD had a lower prevalence of hypertension than ADNI, NACC, and CIMA-Q (χ²=9.33–39.03, *p*<.009). Hypertension did not differ among the other three datasets after correction for multiple comparisons.

Cognitive performance also differed across datasets. MoCA scores differed across all four datasets (*F*=49.73, *p*<.001), with scores increasing from ADNI to NACC, CIMA-Q, and PREVENT-AD (all pairwise *t*>2.36, *p*<.018). Trail Making Test B completion time also differed (*F*=8.93, *p*<.001). NACC participants had longer completion times than participants from ADNI, CIMA-Q, and PREVENT-AD (*t*=3.31–5.26, *p*<.004), whereas the remaining datasets did not differ from one another.

### Demographic differences across prediction horizons

The two, three, four, and five year samples included 265, 208, 195, and 163 participants, respectively. These samples included 43, 60, 86, and 106 converters. Because participants could contribute to more than one horizon, comparisons across horizons are considered descriptive.

Age differed across horizons (*F*=5.60, *p*<.001), with participants in the five-year sample being older than those in the two-year sample (*t*=−3.86, *p*<.001). Trail Making Test B completion time also differed (*F*=6.68, *p*<.001), with longer completion times in the five-year sample than in the two- and three-year samples (*t*=−3.82, *p*=.001 and *t*=−3.03, *p*=.013, respectively). Although the omnibus difference in education was significant (*F*=2.71, *p*=.044), none of the pairwise comparisons remained significant after correction. MoCA scores did not differ across horizons (*F*=2.53, *p*=.056). There were also no differences in sex (χ²=0.06, *p*=.997), APOE4 carrier status (χ²=0.48, *p*=.923), or hypertension (χ²=4.51, *p*=.211).

### Model Results

Performance of the best-performing model for each feature set and prediction horizon is presented in Figure 3 and Table 3. ROC curves for the final best models are presented in Figure 5. Logistic regression was selected as the best model in 8 of 16 horizons by feature-set conditions (Figure 3C). Random forest was selected in 6 conditions. XGBoost was selected in 2 conditions.

**Figure 3.**
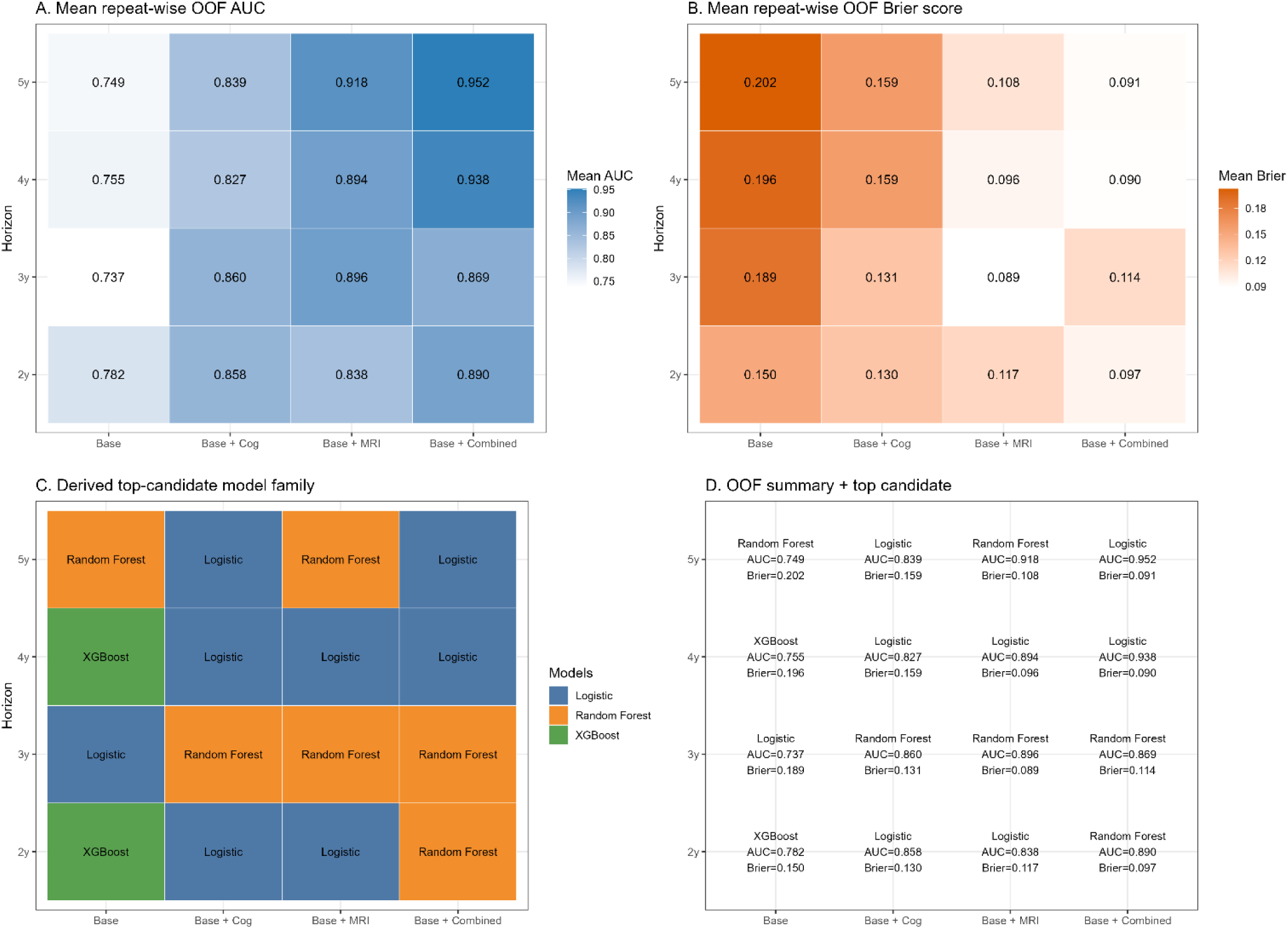
Predictive performance across feature sets. Panel A shows mean out-of-fold (OOF) AUC values for each feature set and prediction horizon. Panel B shows mean OOF Brier scores. Panel C shows the best model type selected for each condition. Panel D summarizes the most frequently selected model and the mean OOF AUC and Brier score for each feature set and horizon.

**Table 1:** Demographic and clinical information across datasets.

| Section | Variable | ADNI | NACC | CIMA-Q | PREVENT-AD |
| --- | --- | --- | --- | --- | --- |
| *Demographics* | N | 270 (36 converters) | 749 (68 converters) | 130 (52 converters) | 203 (39 converters) |
|  | Age (years) | 71.71 (6.16) | 71.05 (9.81) | 73.05 (5.20) | 66.00 (5.36) |
|  | Sex (M:F, %M) | 106:164 (39.26%) | 282:467 (37.65%) | 38:92 (29.23%) | 56:147 (27.59%) |
|  | Education (years) | 16.74 (2.40) | 15.73 (5.44) | 15.45 (3.44) | 15.42 (3.40) |
|  | APOE4 carrier, n (%) | 92 (36.08%) | 227 (34.03%) | 18 (21.43%) | 82 (40.39%) |
|  | Hypertension, n (%) | 106 (39.85%) | 242 (47.36%) | 48 (37.21%) | 43 (21.72%) |
| *Cognition* | MoCA | 25.76 (2.60) | 26.44 (2.62) | 27.69 (1.45) | 28.08 (1.50) |
|  | Trail Making Test B (s) | 79.58 (39.09) | 93.96 (58.49) | 82.42 (30.55) | 78.74 (26.03) |

\*Statistically Significant

**Table 2.** Horizon specific sample information.

| Section | Variable | 2-year | 3-year | 4-year | 5-year |
| --- | --- | --- | --- | --- | --- |
| *Demographics* | N | 265 (43 converters) | 208 (60 converters) | 195 (86 converters) | 163 (106 converters) |
|  | Age (years) | 71.16 (7.43) | 72.20 (7.38) | 72.90 (8.15) | 74.25 (8.39) |
|  | Sex (M:F, %M) | 93:172 (35.09%) | 75:133 (36.06%) | 70:125 (35.90%) | 58:105 (35.58%) |
|  | Education (years) | 16.58 (2.65) | 16.33 (2.69) | 16.06 (2.86) | 15.87 (2.92) |
|  | APOE4 carrier, n (%) | 90 (34.75%) | 65 (32.18%) | 64 (33.68%) | 50 (32.05%) |
|  | Hypertension, n (%) | 86 (34.54%) | 74 (38.74%) | 65 (38.92%) | 59 (45.74%) |
| *Cognition* | MoCA | 26.44 (2.48) | 26.04 (2.55) | 26.08 (2.67) | 25.69 (2.79) |
|  | Trail Making Test B (s) | 81.51 (43.54) | 85.04 (48.35) | 93.94 (59.31) | 103.74 (65.11) |

\*Statistically Significant

**Table 3:**
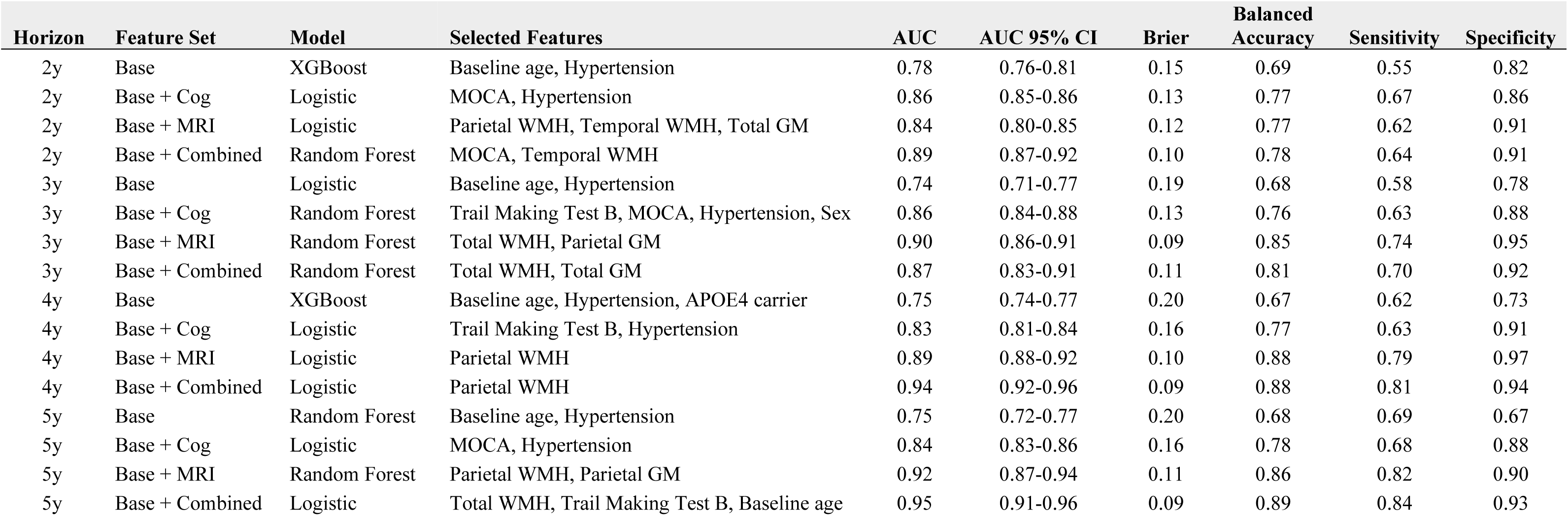
Out-of-fold (OOF) predictive performance across feature sets and prediction horizons.

### Base model

The base feature set (age, sex, education, APOE4 carrier status, and hypertension) achieved mean AUC values between 0.74 and 0.78 across the four horizons (Table 3). At the 2-year horizon, XGBoost achieved the highest AUC of 0.78 (95% CI: 0.76, 0.81; Brier = 0.15). At the 3-year horizon, logistic regression achieved an AUC of 0.74 (95% CI: 0.71, 0.77; Brier = 0.19). At the 4-year horizon, XGBoost achieved an AUC of 0.75 (95% CI: 0.74, 0.77; Brier = 0.20). At the 5-year horizon, random forest achieved an AUC of 0.75 (95% CI: 0.72, 0.77; Brier = 0.20).

### Base and cognition model

Adding MoCA and TMT-B to the base features improved performance at all horizons (Table 3, Figure 4). At the 2-year horizon, logistic regression achieved an AUC of 0.86 (95% CI: 0.85, 0.86; Brier = 0.13; ΔAUC = +0.08 vs. base). At the 3-year horizon, random forest achieved an AUC of 0.86 (95% CI: 0.84, 0.88; Brier = 0.13; ΔAUC = +0.12). At the 4-year horizon, logistic regression achieved an AUC of 0.83 (95% CI: 0.81, 0.84; Brier = 0.16; ΔAUC = +0.08). At the 5-year horizon, logistic regression achieved an AUC of 0.84 (95% CI: 0.83, 0.86; Brier = 0.16; ΔAUC = +0.09).

**Figure 4.**
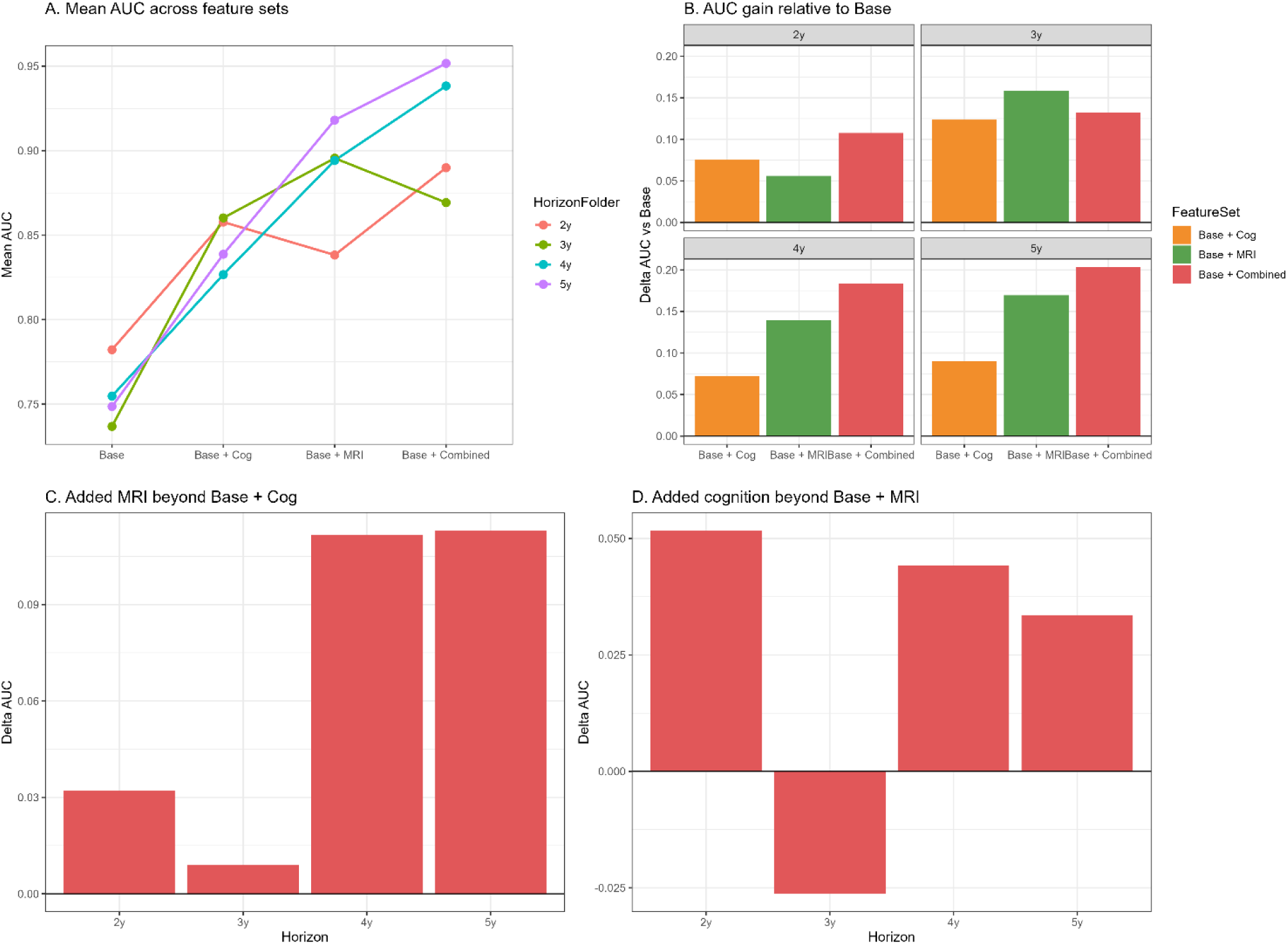
Incremental predictive value of cognition and MRI features. Panel A shows mean OOF AUC across feature sets and horizons. Panel B shows the AUC gain for each feature set compared with the base model. Panel C shows the added value of MRI beyond the base plus cognition model. Panel D shows the added value of cognition beyond the base plus MRI model.

**Figure 5.**
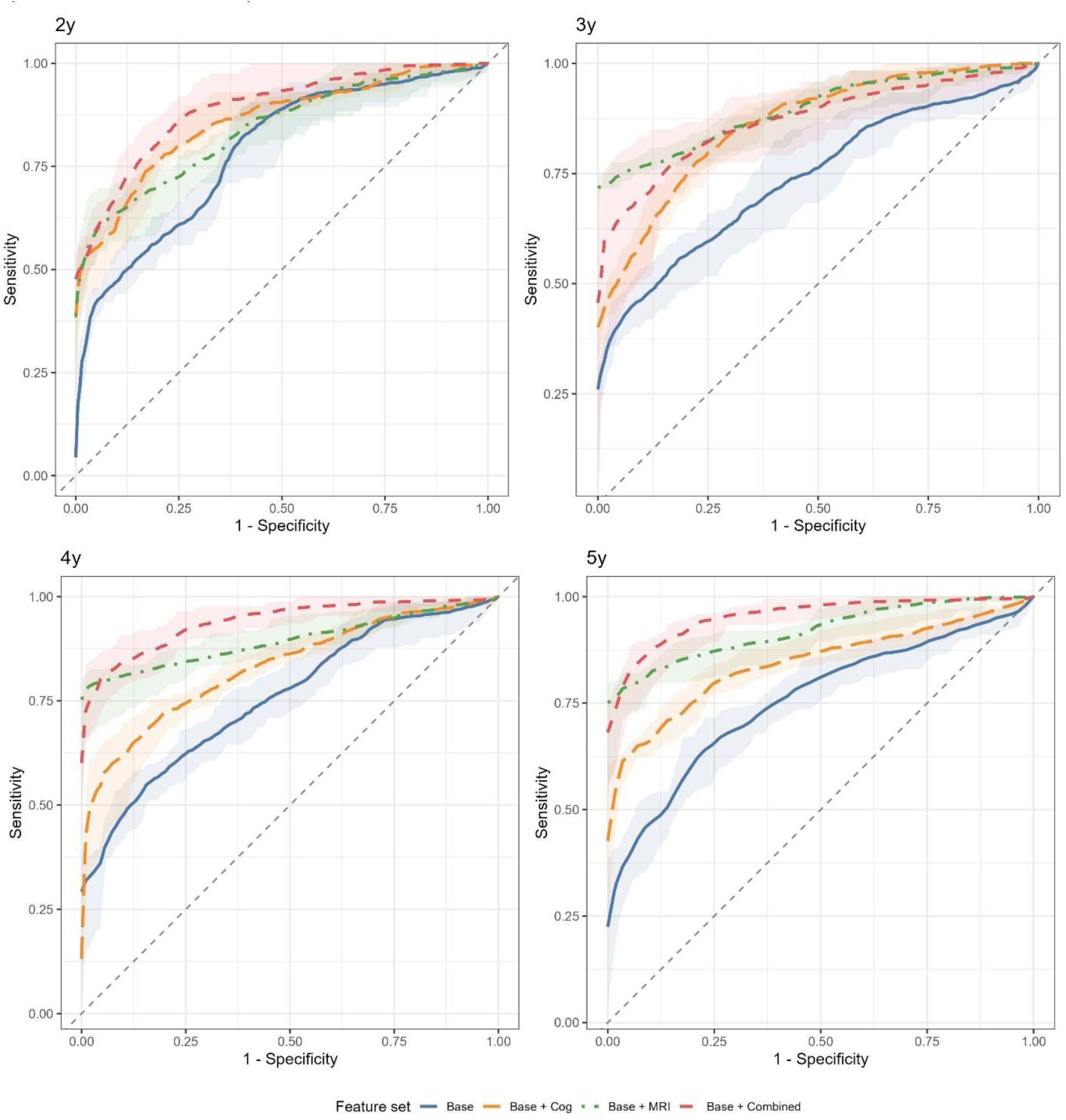
Mean out-of-fold ROC curves across repeated cross-validation. ROC curves show model discrimination across the 2, 3, 4, and 5-year prediction horizons. Each curve represents the mean of the 20 outer repeats OOF ROC curves, with shaded areas showing the empirical 2.5th–97.5th percentile range across repeats. Curves farther from the diagonal line indicate better prediction.

### Base and MRI model

Adding regional WMH and GM to the base features improved performance at all horizons and yielded the highest AUC at the 3-year horizon (Table 3, Figure 3A). At the 2-year horizon, logistic regression achieved an AUC of 0.84 (95% CI: 0.80, 0.85; Brier = 0.12; ΔAUC = +0.06 vs. base). At the 3-year horizon, random forest achieved an AUC of 0.90 (95% CI: 0.86, 0.91; Brier = 0.09; ΔAUC = +0.16). At the 4-year horizon, logistic regression achieved an AUC of 0.89 (95% CI: 0.88, 0.92; Brier = 0.10; ΔAUC = +0.14). At the 5-year horizon, random forest achieved an AUC of 0.92 (95% CI: 0.87, 0.94; Brier = 0.11; ΔAUC = +0.17).

### Combined model

Including all base, cognitive, and MRI features achieved the highest AUC at the 2, 4, and 5 year horizons (Table 3). At the 2-year horizon, random forest achieved an AUC of 0.89 (95% CI: 0.87, 0.92; Brier = 0.10; ΔAUC = +0.11 vs. base). At the 3-year horizon, random forest achieved an AUC of 0.87 (95% CI: 0.83, 0.91; Brier = 0.11; ΔAUC = +0.13). At the 4-year horizon, logistic regression achieved an AUC of 0.94 (95% CI: 0.92, 0.96; Brier = 0.09; ΔAUC = +0.19). At the 5-year horizon, logistic regression achieved the highest AUC across all conditions: 0.95 (95% CI: 0.91, 0.96; Brier = 0.09; ΔAUC = +0.20). At the 3-year horizon, the combined set did not outperform the base and MRI set.

### Incremental predictive value

The incremental value of each feature set relative to the base model is presented in Figure 4. Both cognition and MRI features improved prediction beyond base features at all horizons. The MRI set produced larger AUC gains than the cognition set at the 3-year (+0.16 vs. +0.12), 4-year (+0.14 vs. +0.07), and 5-year (+0.17 vs. +0.09) horizons (Figure 4B). At the 2-year horizon, cognition produced a larger gain than MRI (+0.08 vs. +0.06).

Adding MRI features to the base and cognition set (combined vs. base and cognition) yielded AUC gains at all horizons: +0.03 at 2 years, +0.01 at 3 years, +0.11 at 4 years, and +0.11 at 5 years (Figure 4C). Adding cognitive features to the base and MRI set (combined vs. base and MRI) improved AUC at the 2-year (+0.05), 4-year (+0.04), and 5-year (+0.03) horizons but reduced AUC at the 3-year horizon (−0.03) (Figure 4D). These are descriptive comparisons of repeat averaged OOF performance.

### Feature selection

Feature selection across the outer repeats is presented in Figure 6A. Within the base set, baseline age and hypertension formed the most frequently selected winning subset at every horizon, accounting for 16 of 20 repeats at 2 years, 11 of 20 at 3 years, 11 of 20 at 4 years, and 15 of 20 at 5 years. Within the base and cognition set, MoCA and hypertension formed the leading subset at 2 years (11 of 20 repeats). At 3 years, the leading subset included TMT-B, MoCA, hypertension, and sex (7 of 20 repeats). At 4 years, TMT-B and hypertension formed the leading subset (7 of 20 repeats). At 5 years, two subsets tied at 6 of 20 repeats each: TMT-B, MoCA, and hypertension, and the same three features with APOE4 carrier status added.

**Figure 6.**
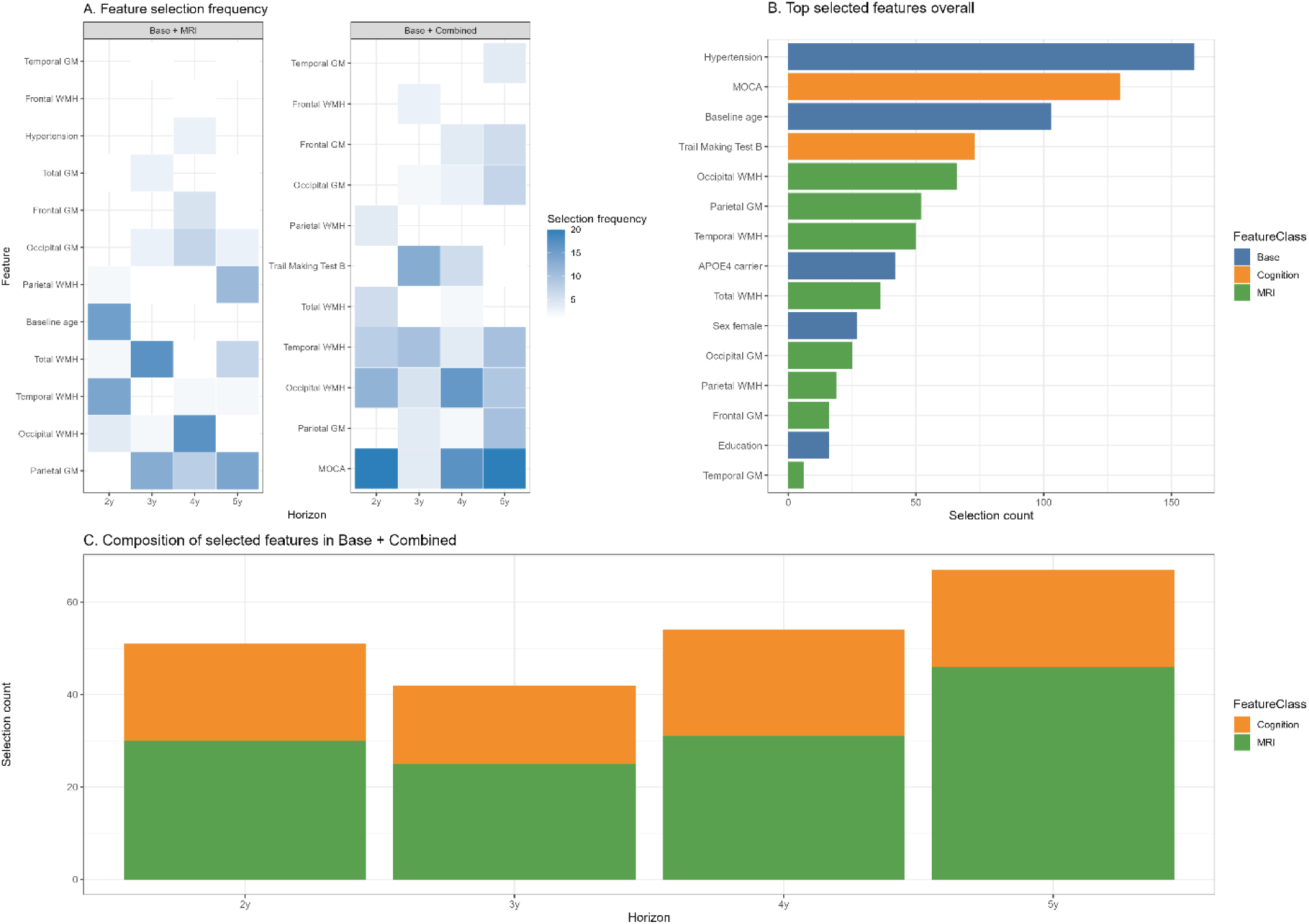
Feature selection results. Panel A shows how often each feature was selected across horizons in the base plus MRI and combined feature sets. Panel B shows the most frequently selected features overall. Panel C shows the contribution of base, cognition, and MRI features among selected features in the combined model.

Within the base and MRI set, the leading subset changed across horizons: temporal WMH and baseline age at 2 years (11 of 20 repeats), total WMH and parietal GM at 3 years (10 of 20), occipital WMH and occipital GM at 4 years (6 of 20), and parietal WMH and parietal GM at 5 years (9 of 20). MRI features therefore accounted for 50% of the leading subset at 2 years and 100% at the 3-, 4-, and 5-year horizons. Within the combined set, the leading subsets paired cognitive and MRI features at each horizon: MoCA and occipital WMH at 2 years (9 of 20 repeats), TMT-B and temporal WMH at 3 years (5 of 20), MoCA and occipital WMH at 4 years (7 of 20), and temporal WMH, MoCA, and occipital GM at 5 years (4 of 20). No base feature entered a winning subset in the combined set at any horizon, where MRI features accounted for 57% to 69% of selections across horizons (Figure 6C).

### Predicted risk distributions

Predicted probability distributions based on each participant’s mean OOF predicted probability across the 20 repetitions are presented in Figure 7A, B, and D. Across all horizons and feature sets, converters had higher mean predicted probabilities than non-converters. The largest differences between converters and non-converters were observed for the combined set at the 2 and 4-year horizons and for the base and MRI set at the 3 and 5-year horizons. In the base and MRI set at the 5-year horizon, converters had a mean predicted probability of 0.83 (SD = 0.28), compared with 0.22 (SD = 0.14) for non-converters. Calibration plots (Figure 7C) indicated closer alignment between predicted probabilities and observed conversion rates for the MRI-containing models at the longer prediction horizons, particularly at 5 years.

**Figure 7.**
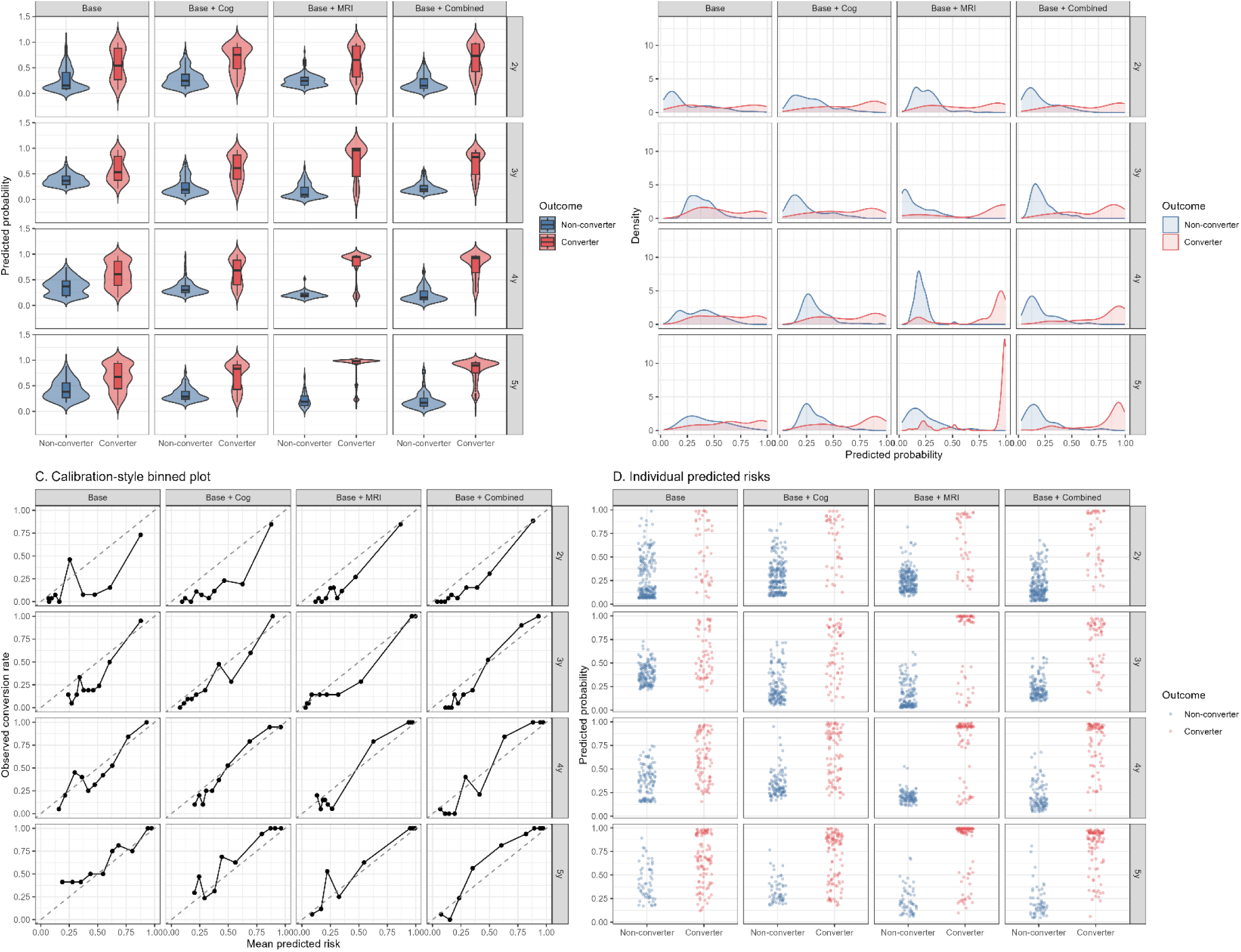
Predicted risk distributions. Rows correspond to the 2, 3, 4, and 5-year prediction horizons, and columns correspond to the base, base and cognition, base and MRI, and combined feature sets. Panel A shows the distributions of predicted probabilities for converters and non-converters using violin and box plots. Panel B shows the corresponding probability density distributions. Panel C shows calibration-style plots comparing mean predicted risk with the observed conversion rate across probability bins. Black points represent the individual bins, the dashed diagonal line represents ideal agreement between predicted and observed risk, and the blue line represents the fitted relationship. Panel D shows participant-level predicted probabilities for converters and non-converters. Across panels, converters generally had higher predicted probabilities than non-converters, with greater separation at the longer prediction horizons. The clearest separation was observed for the base and MRI feature set at the 5-year horizon, whereas greater overlap was generally present at the shorter horizons and for the base feature set.

## Discussion

SCD may reflect early brain changes associated with later cognitive decline, but not all individuals with SCD progress to MCI. Although demographic, genetic, vascular, cognitive, and MRI markers have been associated with conversion, the additional predictive value of regional WMH burden and GM atrophy remains uncertain. Across the four datasets, approximately 15% of participants converted from SCD to MCI. This proportion is consistent with previous reviews and studies although estimates may vary depending on follow-up duration, sample characteristics, and the definition of SCD [52,53]. It is also unclear whether cognitive and MRI predictors differ in their usefulness across shorter and longer prediction horizons. The present study addressed these gaps by comparing different combinations of predictors for SCD to MCI conversion across 2, 3, 4, and 5-year horizons. The base feature set (i.e., baseline age, sex, education, APOE4 carrier status, and hypertension) and base plus MRI feature set (i.e., regional WMH and GM) achieved the highest predictive performance (as measured by AUC) at the 3, 4, and 5-year horizons. The combined feature set (i.e., base, cognitive, and MRI features) achieved the highest predictive performance, as measured by AUC, at the 2, 4, and 5-year horizons. The base plus MRI feature set achieved the highest AUC at the 3-year horizon, where it outperformed the combined feature set. These findings are consistent with previous studies which have similarly shown that clinical and MRI features can predict future conversion to dementia [13,54,55]. In our previous study using the ADNI and NACC datasets to predict post-mortem neuropathology, models that included MRI features achieved higher accuracy than models with clinical features alone, and MRI predictors became more informative as the time between data collection and death increased [13].

Both cognitive and MRI features improved prediction performance in contrast to the base model across all prediction horizons. Nonetheless, the performance of each feature set was dependent on the prediction horizon. For example, at the 2-year horizon, adding cognitive features led to a larger increase in AUC than adding MRI features. In contrast, adding MRI features produced larger increases in AUC than adding cognitive features at the 3, 4, and 5-year horizons. This difference was largest at the 5-year horizon. Subtle cognitive differences measured using the MoCA and TMT-B may identify individuals who are already approaching the MCI threshold and are therefore more likely to convert within a shorter timeframe.

Adding cognition (e.g., MoCA) to the base and MRI set improved AUC at the 2, 4, and 5-year horizons but reduced AUC at the 3-year horizon. The improvements at 4 and 5 years were modest relative to the gains obtained by adding MRI to the base and cognition set. Prior work suggests that including too many predictors in modest to small sample sizes can reduce generalization [48, 56]. Our nested cross-validation procedure, which favors parsimonious subsets, mitigated this risk, yet the cognitive features offered little incremental value beyond MRI (regional WMH and GM), implying that the cognitive information was largely redundant with the MRI features. One possible explanation is that WMH accumulation and GM atrophy may emerge years before cognitive decline can be detected using standard cognitive assessments [2, 5]. Given that AD-related structural brain changes are known to accumulate 10 to 20 years before clinical diagnosis [1, 2], MRI features may capture early pathology that is not yet reflected in standard cognitive tests.

Regional WMH features were consistently represented in the feature selection results. In the final models, parietal WMH was selected at the 2, 4, and 5-year horizons for the base plus MRI feature set. At the 4-year horizon, parietal WMH was the only feature retained in both the base plus MRI and combined models. Total WMH was selected at the 3 and 5 year horizons. Across the outer repeats, temporal and occipital WMH also appeared in leading subsets, indicating that predictive information was not limited to parietal or total WMH burden. Parietal WMH progression has been shown to predict AD in older adults [57], and parietal white matter lesions in AD have been associated with cortical neurodegenerative pathology [58, 59]. Regional WMH burden has also been associated with specific cognitive modalities. For instance, posterior WMHs have been associated with memory decline [4, 60]. In our previous study, we observed that temporal and occipital WMHs showed the strongest cognitive associations in older adults with MCI [4]. Present results which also show frequent selection of temporal and occipital WMH across repeats align with our previous findings. GM regions, particularly parietal and total GM, contributed to improved prediction at several horizons. Together these findings suggest that regional WMH burden and concurrent GM atrophy provide complementary information for long-term prediction of conversion from SCD to MCI.

Logistic regression was found to be the best performing ML model across horizons compared to random forest and XGBoost. The frequent selection of logistic regression may indicate that the relationships between the selected features and conversion were often linear. However, nonlinear classifiers also contributed. This finding differs from our previous AD neuropathology prediction study, in which nonlinear complex models, including gradient boosting and SVM, outperformed simpler linear regression models [13]. The neuropathology study involved continuous outcomes across a wider range of pathological severity, whereas the present study involved a binary classification task with a smaller feature set. In neuroimaging studies with limited samples, simpler models often generalize better than complex models [26, 48].

Studies examining SCD to MCI have highlighted MoCA scores, stroke history, amygdala volume, and WMH burden as strong predictors [22, 27, 61, 62]. Most of these studies relied on a single follow-up window or evaluated cognition and neuroimaging in separate models. By pooling four independent cohorts and testing four predefined feature sets across four horizons, the present study allowed the incremental value of cognitive and MRI variables to be examined as a function of time.

Several limitations warrant mention. First, SCD was defined differently across datasets. Although each definition required a self-reported concern without objective impairment, item wording and thresholds varied, potentially influencing baseline characteristics and conversion rates. Second, sample sizes declined at longer horizons; only 163 participants contributed data at five years. Although nested cross-validation helped curb over-fitting, estimates at longer horizons are inevitably less stable. Third, consistent fluid biomarkers were unavailable across datasets. Cerebrospinal fluid or plasma measures of amyloid and tau might further stratify risk and improve prediction of older adults on the AD trajectory [1, 63]. Combining MRI features with fluid biomarkers should be examined in future studies. Similarly, measures of depression and anxiety were also not included. There is evidence to suggest that both measures influence SCD trajectories and conversion [27].

Overall, the present study showed that cognitive and MRI features improved the prediction of SCD to MCI conversion beyond demographic, genetic, and vascular risk factors. Cognitive features provided greater benefit at the shorter prediction horizon, while regional WMH and GM measures were more informative at longer horizons. Regional WMH features were consistently represented, with parietal WMH often selected in several final models. These findings indicate that the predictors of SCD conversion may differ depending on the timeframe being examined and support the inclusion of regional MRI measures in models of longer-term risk.

## Data Availability

Data used in preparation of this article were obtained from the PRe-symptomatic EValuation of Experimental or Novel Treatments for Alzheimers Disease (PREVENT-AD) program data release 8.0 (https://www.centrestopad.com/).
The CIMA-Q data are available to the research community to address research questions related to dementia and aging, upon approval by the User Access Committee. Access is limited to CIMA-Q members. However, membership is open to all scientists interested in dementia research. Detailed information on the procedure for submitting a data access request can be found on the CIMA-Q website (https://www.cima-q.ca/en/home/).
Data used in preparation of this article were also obtained from the National Alzheimers Coordinating Center (NACC- https://naccdata.org/) database, including the NACC Uniform Data Set (UDS), and MRI Data Set (Beekly et al., 2004; Besser, Kukull, Knopman, et al., 2018; Besser, Kukull, Teylan, et al., 2018). The image processing and WMH segmentation pipelines used are open source and available at https://github.com/VANDAlab/Preprocessing_Pipeline. The lobar atlas used to derive regional WMH metrics is also available at https://zenodo.org/records/7930159.
The data used in this study were obtained from the Alzheimers Disease Neuroimaging Initiative (ADNI) database (adni.loni.usc.edu). ADNI was launched in 2003 as a public-private partnership, led by Principal Investigator Michael W. Weiner, MD. The image processing and WMH segmentation pipelines used are open source and available at https://github.com/VANDAlab/Preprocessing_Pipeline. MRI protocols and imaging parameters can be found at: http://adni.loni.usc.edu/methods/mri-tool/mri-analysis/. All participant MRI data (baseline and longitudinal) were downloaded from the ADNI public website.

## Acknowledgments

Data collection and sharing for this project was funded by the Alzheimer’s Disease Neuroimaging Initiative (ADNI) (National Institutes of Health Grant U01 AG024904) and DOD ADNI (Department of Defense award number W81XWH-12-2-0012). ADNI is funded by the National Institute on Aging, the National Institute of Biomedical Imaging and Bioengineering, and through generous contributions from the following: AbbVie, Alzheimer’s Association; Alzheimer’s Drug Discovery Foundation; Araclon Biotech; BioClinica, Inc.; Biogen; Bristol-Myers Squibb Company; CereSpir, Inc.; Cogstate; Eisai Inc.; Elan Pharmaceuticals, Inc.; Eli Lilly and Company; EuroImmun; F. Hoffmann-La Roche Ltd and its affiliated company Genentech, Inc.; Fujirebio; GE Healthcare; IXICO Ltd.; Janssen Alzheimer Immunotherapy Research & Development, LLC.; Johnson & Johnson Pharmaceutical Research & Development LLC.; Lumosity; Lundbeck; Merck & Co., Inc.; Meso Scale Diagnostics, LLC.; NeuroRx Research; Neurotrack Technologies; Novartis Pharmaceuticals Corporation; Pfizer Inc.; Piramal Imaging; Servier; Takeda Pharmaceutical Company; and Transition Therapeutics. The Canadian Institutes of Health Research is providing funds to support ADNI clinical sites in Canada. Private sector contributions are facilitated by the Foundation for the National Institutes of Health (www.fnih.org). The grantee organization is the Northern California Institute for Research and Education, and the study is coordinated by the Alzheimer’s Therapeutic Research Institute at the University of Southern California. ADNI data are disseminated by the Laboratory for Neuro Imaging at the University of Southern California.

Data used in the preparation of this article were obtained from the PRe-symptomatic EValuation of Experimental or Novel Treatments for Alzheimer’s Disease (PREVENT-AD) program data release 8.0. PREVENT-AD was launched in 2011 as a $13.5 million, 7-year public-private partnership using funds provided by McGill University, the Fonds de Recherche du Québec – Santé (FRQ-356162), an unrestricted research grant from Pfizer Canada, the J.L. Levesque Foundation, the Douglas Hospital Research Centre and Foundation, the Government of Canada, the Canada Fund for Innovation, the Canadian Institutes of Health Research (SV: 178385, JP: 153287, 178210, LC: 165921, TS: 175328,) the Alzheimer Society of Canada, the National Institutes of Health of the United States (NS: NIH AG068563), the Alzheimer Association (SB: AARG-NTF 926696) and Brain Canada Foundation. Parts of the data used in this article were obtained from the Consortium for the Early Identification of Alzheimer’s disease (CIMA-Q; https://www.cima-q.ca/en/home/). The CIMA-Q investigators contributed to the design, protocols and implementation of the study, as well as the collection of clinical, cognitive and neuroimaging data and biological samples. A complete list of the CIMA-Q investigators can be found at https://www.cima-q.ca/en/home/. The CIMA-Q is supported by the Fonds de recherche du Québec–Santé (FRQS) - Pfizer Innovation Program, FRQ cohort funds, the Quebec Network for Research on aging, the Fondation Courtois (NeuroMod project), the Consortium for the Neurodegeneration associated with Aging, and the Fondation Famille Lemaire. This study was supported by research funds from the Canadian Institutes of Health Research (CIHR).

The authors acknowledge use of Compute Canada (https://alliancecan.ca/en) resources for performing the image processing and WMH segmentations in the presented work. The NACC database is funded by NIA/NIH Grant U24 AG072122. NACC data are contributed by the NIA-funded ADRCs: P30 AG062429 (PI James Brewer, MD, PhD), P30 AG066468 (PI Oscar Lopez, MD), P30 AG062421 (PI Bradley Hyman, MD, PhD), P30 AG066509 (PI Thomas Grabowski, MD), P30 AG066514 (PI Mary Sano, PhD), P30 AG066530 (PI Helena Chui, MD), P30 AG066507 (PI Marilyn Albert, PhD), P30 AG066444 (PI David Holtzman, MD), P30 AG066518 (PI Lisa Silbert, MD, MCR), P30 AG066512 (PI Thomas Wisniewski, MD), P30 AG066462 (PI Scott Small, MD), P30 AG072979 (PI David Wolk, MD), P30 AG072972 (PI Charles DeCarli, MD), P30 AG072976 (PI Andrew Saykin, PsyD), P30 AG072975 (PI Julie A. Schneider, MD, MS), P30 AG072978 (PI Ann McKee, MD), P30 AG072977 (PI Robert Vassar, PhD), P30 AG066519 (PI Frank LaFerla, PhD), P30 AG062677 (PI Ronald Petersen, MD, PhD), P30 AG079280 (PI Jessica Langbaum, PhD), P30 AG062422 (PI Gil Rabinovici, MD), P30 AG066511 (PI Allan Levey, MD, PhD), P30 AG072946 (PI Linda Van Eldik, PhD), P30 AG062715 (PI Sanjay Asthana, MD, FRCP), P30 AG072973 (PI Russell Swerdlow, MD), P30 AG066506 (PI Glenn Smith, PhD, ABPP), P30 AG066508 (PI Stephen Strittmatter, MD, PhD), P30 AG066515 (PI Victor Henderson, MD, MS), P30 AG072947 (PI Suzanne Craft, PhD), P30 AG072931 (PI Henry Paulson, MD, PhD), P30 AG066546 (PI Sudha Seshadri, MD), P30 AG086401 (PI Erik Roberson, MD, PhD), P30 AG086404 (PI Gary Rosenberg, MD), P20 AG068082 (PI Angela Jefferson, PhD), P30 AG072958 (PI Heather Whitson, MD), P30 AG072959 (PI James Leverenz, MD).

## Consent Statement

Written informed consent was obtained from participants or their study partner

## Availability of data and materials

The data used in this study were obtained from the Alzheimer’s Disease Neuroimaging Initiative (ADNI) database (adni.loni.usc.edu). ADNI was launched in 2003 as a public-private partnership, led by Principal Investigator Michael W. Weiner, MD. The image processing and WMH segmentation pipelines used are open source and available at https://github.com/VANDAlab/Preprocessing_Pipeline. MRI protocols and imaging parameters can be found at: http://adni.loni.usc.edu/methods/mri-tool/mri-analysis/. All participant MRI data (baseline and longitudinal) were downloaded from the ADNI public website.

Data used in preparation of this article were also obtained from the National Alzheimer’s Coordinating Center (NACC, https://naccdata.org/) database, including the NACC Uniform Data Set (UDS), and MRI Data Set (Beekly et al., 2004; Besser, Kukull, Knopman, et al., 2018; Besser, Kukull, Teylan, et al., 2018). The image processing and WMH segmentation pipelines used are open source and available at https://github.com/VANDAlab/Preprocessing_Pipeline. The lobar atlas used to derive regional WMH metrics is also available at https://zenodo.org/records/7930159.

Data used in preparation of this article were obtained from the PRe-symptomatic EValuation of Experimental or Novel Treatments for Alzheimer’s Disease (PREVENT-AD) program, data release 8.0 (https://www.centrestopad.com/).

The CIMA-Q data are available to the research community to address research questions related to dementia and aging, upon approval by the User Access Committee. Access is limited to CIMA-Q members. However, membership is open to all scientists interested in dementia research. Detailed information on the procedure for submitting a data access request can be found on the CIMA-Q website (https://www.cima-q.ca/en/home/).

## Consent for publication

Not applicable.

